# Clinical characteristics of 4490 COVID-19 patients in Africa: A meta-analysis

**DOI:** 10.1101/2020.10.20.20215905

**Authors:** Testimony Jesupamilerin Olumade, Leonard Ighodalo Uzairue

## Abstract

**Introduction:** The novel coronavirus disease-2019 (COVID-19) pandemic that started in December 2019 has affected over 39 million people and killed over 1.1 million people. While more studies are published to help us understand the virus, there is a dearth of studies on the clinical characteristics and associated outcomes of the Severe Acute Respiratory Syndrome Coronavirus 2 (SARS-CoV-2) on the African continent.

**Methods:** We evaluated evidence from previous studies in Africa available in six databases between January 1 and October 6, 2020. Meta-analysis was then performed using Open-Meta Analyst software.

**Results:** A total of seven studies including 4490 COVID-19 patients were included. The result of the meta-analysis showed 68.8% of infected patients were male. Common symptoms presented (with their incidences) were fever (42.8%), cough (33.3%), headache (11.3%), breathing problems (16.8%). Other minor occurring symptoms included diarrhea (7.5%), and rhinorrhea (9.4%). Fatality rate was 5.6%.

**Conclusion:** This study presents the first description and analysis of the clinical characteristics of COVID-19 patients in Africa. The most common symptoms are fever, cough and breathing problems.

## INTRODUCTION

A disease that was first reported in early December 2019^1^ has led to a pandemic in 2020 that has resulted in about 39 million cases and over 1.1 million deaths as of 18th October, 2020^2^. Nearly every continent of the earth, except Antarctica, has felt the impact of the novel Coronavirus Disease 2019 (COVID-19) caused by the Severe Acute Respiratory Syndrome Coronavirus 2 (SARS-CoV-2)^3,4^. It was predicted that Africa will be the worst hit with the pandemic, having no less than 223 million cases and more than 150,000 fatalities^3,5^. However, eight months into the pandemic, this is not the case. The WHO African Region, comprising only 47 member states on the continent (without Djibouti, Egypt, Libya, Morocco, Somalia, Sudan and Tunisia), has reported only 3.2% of all COVID-19 cases (1,259,192 out of 39,596,858) and 2.6% of deaths (28,313 out of 1,107,374) globally as of October 18^th^, 2020^2,6^.

Nevertheless, to improve our understanding and management of this novel disease, it remains pertinent to define and analyze the clinical characteristics and outcomes of the disease in patients^7^. Several studies have described the clinical and epidemiological characteristics, risk factors, case management, and associated outcomes of different patient cohorts with COVID- 19 globally^1,7–15^. Clinical features, diagnosis and treatment of COVID-19 have also been reviewed elsewhere^16^. However, there is limited information on the clinical characteristics of COVID-19 in patients in Africa. In this article, we describe a meta-analysis of the clinical characteristics of COVID-19 patients in Africa.

## METHODOLOGY

### Search databases and search strategyy

PubMed, Google scholar, Scopus, The Cochrane Library, EMBASE, and Africa journal online (AJOL) were electronically searched to collect clinical studies on the clinical characteristics of COVID-19 from January 1, 2020 to October 6, 2020. We also performed a manual search of the reference lists of included studies to avoid omitting any eligible study. Only studies written in English language and published online were included. The following terms were used in search alone OR in combination: “Coronavirus”OR“2019-nCoV”OR“COVID-19”OR“SARS-CoV-2” AND “Africa” OR “Comoros” OR “Djibouti” OR “Madagascar” OR “Malawi” OR “Seychelles” OR “Cameroon” OR “Central African Republic” OR “Chad” OR “Congo” OR “Equatorial Guinea” OR “Atlantic Islands” OR “Gabon” OR “Morocco” OR “South Sudan” OR “Sudan” OR “Botswana” OR “Lesotho” OR “Swaziland” OR “Benin” OR “Burkina Faso” OR “Cape Verde” OR “Ghana” OR “Guinea” OR “Guinea-Bissau” OR “Mauritania” OR “Niger” OR “Senegal” OR “Sierra Leone” OR “Togo” OR “Burundi” OR “Eritrea” OR “Ethiopia” OR “Kenya” OR “Mozambique” OR “Rwanda” OR “Somalia” OR “Tanzania” OR “Uganda” OR “Zambia” OR “Zimbabwe” OR “Angola” OR “Algeria” OR “Egypt” OR “Tunisia” OR “Namibia” OR “South Africa” OR “Gambia” OR “Liberia” OR “Mali” OR “Nigeria” AND English[lang]. Figure 1 shows the flow chart of the literature screening process.

**Figure 1:**
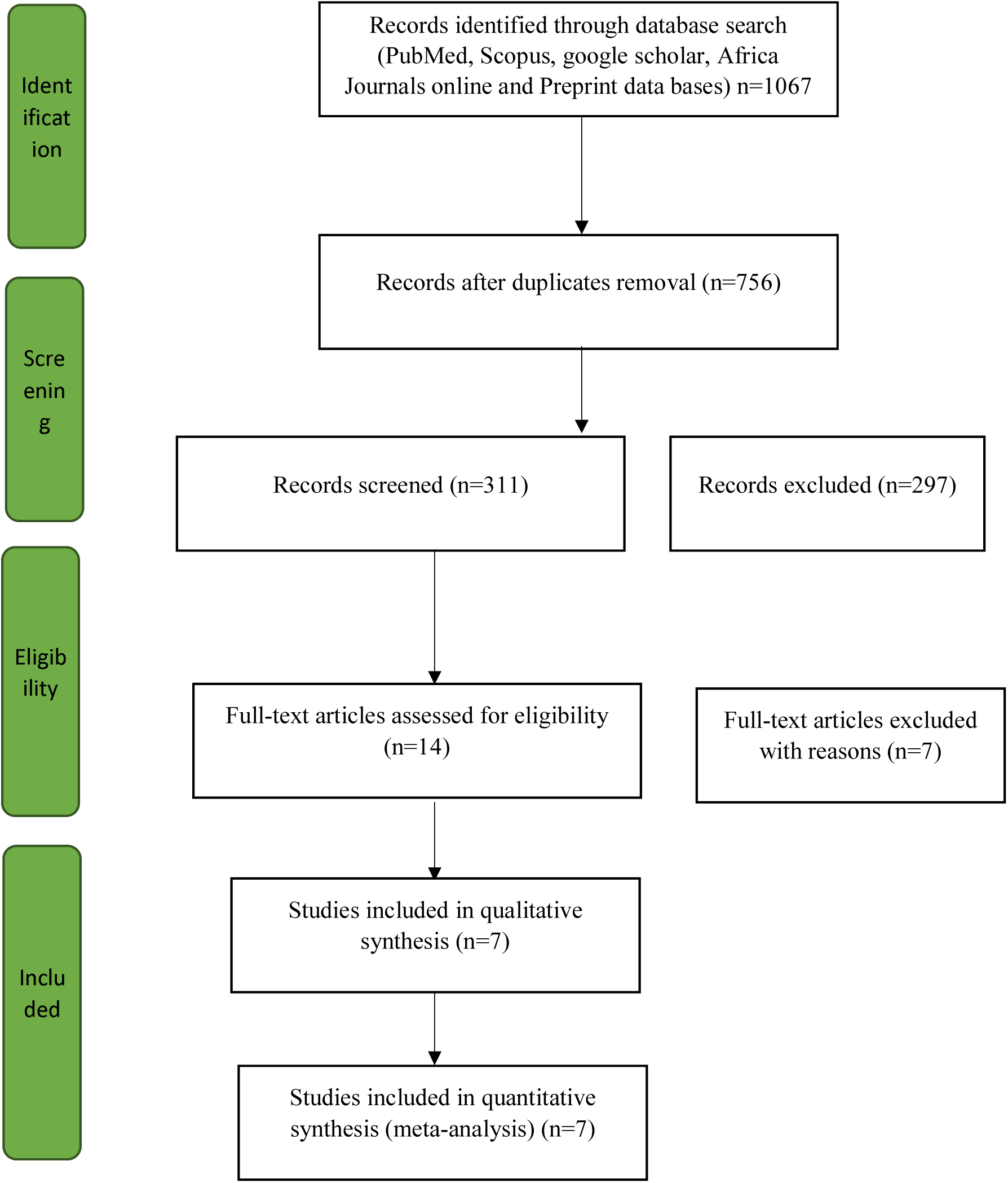
Flow chart of the literature screening process.

### Inclusion and exclusion criteria

The inclusion criteria were as follows: (1) cohort studies, case-control studies, and case series studies; (2) the study population included individuals diagnosed with COVID-19; (3) the primary outcomes were: clinical symptoms, signs, demographics and fatality rate, etc. The exclusion criteria were as follows: (1) overlapping or duplicate studies; (2) The epidemiological analysis with only secondary outcomes such as fatality rate, without the primary outcomes; (3) had no clinical indicators or lacking necessary data.

### Data extraction and quality assessment

Two reviewers, using the inclusion and exclusion criteria, independently selected the literature, and extracted data to an Excel database and any disagreement was resolved by consensus. Data extraction included the first author ‘s surname and the date of publication of the article, study region/country, study design, sample size, age and outcome measurement data such as clinical symptoms. The included studies of this meta-analyses were observational case series studies, so the British National Institute for Clinical Excellence (BNICE) was used to evaluate the study quality by the independent reviewers. The evaluation included 8 items and the total score was 8. Studies with a score greater than 4 were seen as high-quality.

### Statistical analyses

All the meta-analyses were performed by using Open-Meta Analyst^17^ Single-arm meta-analyses were carried out. The heterogeneity was quantified using the I^2^ statistic. The random model was utilized for statistical heterogeneity between the results of each study.

## RESULTS

### Literature retrieval and articles characteristics

A total of 1065 Records were identified during literature retrieval from databases. A total of 7 studies involving 4490 COVID-19 patients were included in this meta-analysis^8,9,15,18–21^ (Figure 1 and Table 1). All studies included in this meta-analysis were conducted in African countries. The characteristics of the studies included in this meta-analysis were published between June 1, 2020 to October 2, 2020. The quality score of the included studies ranged from 5 to 7 on a maximum scale of quality score of 8 using the NICE criteria^22^ (Table 1).

**Table 1:** Basic characteristics and quality score of included studies

| S/N | Studies | Countries | Number of patients | Quality Score according to NICE Criteria | Date Published |
| --- | --- | --- | --- | --- | --- |
| 1 | Chekhlabi *et al.* | Morocco | 15 | 6 | June 1, 2020 |
| 2 | Abimbola-Bowale *et al.* | Nigeria | 32 | 5 | May 6, 2020 |
| 3 | Abdoul Aziz-Diouf *et al.* | Senegal | 9 | 5 |  |
| 4 | Nachega *et al.* | Congo | 766 | 6 | October 2, 2020 |
| 5 | Kirenga *et al.* | Uganda | 56 | 7 | August 24, 2020 |
| 6 | Elimian *et al.* | Nigeria | 3467 | 6 | August 26, 2020 |
| 7 | Otuonye *et al* (Pre-Print) | Nigeria | 154 | 5 | September 24, 2020 |
| **Total** |  |  | **4490** |  |  |

### Gender distribution

The random effect model was used in the meta-analysis. Gender distribution showed that the proportion of male was 68.8% (95% CI, 64.6%-72.9%) with significant heterogeneity I^2^ = 65.16%, p = 0.014 (Figure 2).

**Figure 2:**
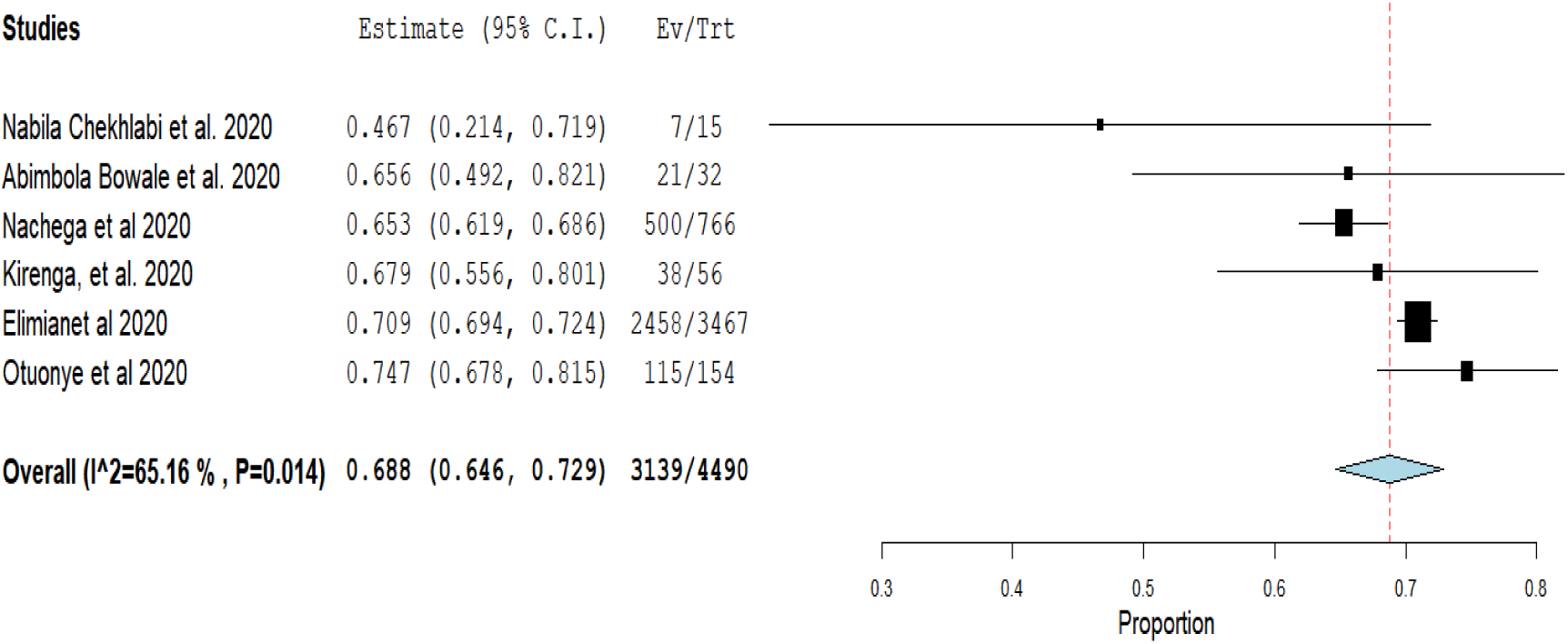
Proportion of males in COVID-19 patients.

### Clinical symptoms

The incidence of commonly encountered symptoms were as follows: Fever (42.8%, 95% CL 30.7%-54.9%), Cough (33.3%, 95% CL, 24.0%-42.5%), Headache (11.3%, 95%CL, 2.9%-25.4%), Breathing problem (16.8%, 95% CL, 4.5%-29.1%) Diarrhea (7.5%, 95%CL, 2.8%-12.2%) and Rhinorrhea (9.4%, 95% CL, 8.5%-10.4%) as show in Table 2.

**Table 2:**
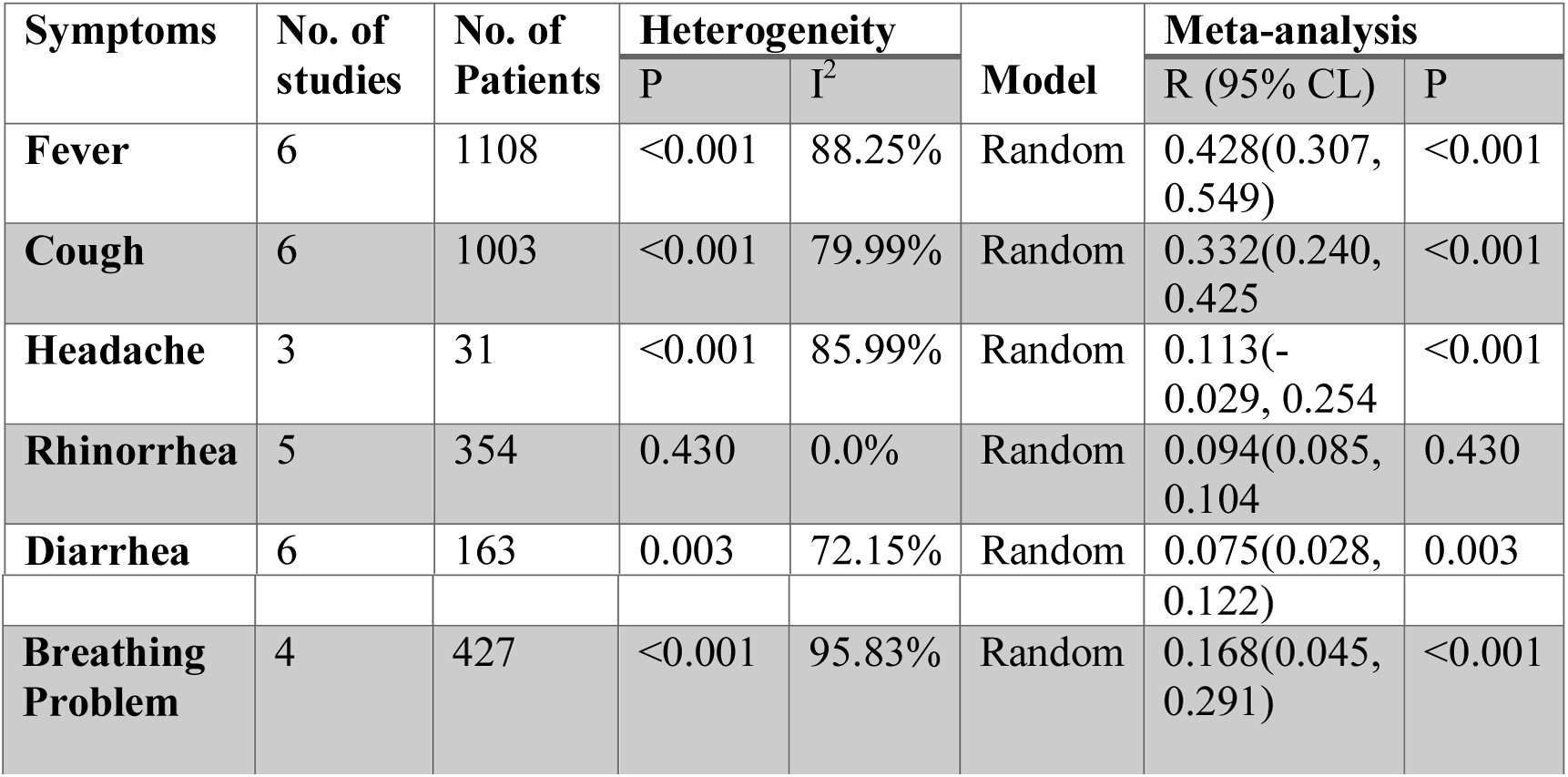
Meta-analysis of different clinical symptoms in COVID-19 patients

### Fatality rate

The fatality rate in the 4490 patients included in the meta-analysis was 5.6% (95% CL, 2.7%- 8.6%) with significant heterogeneity (I^2^=85.9%, p= <0.001) (Figure 3).

**Figure 3:**
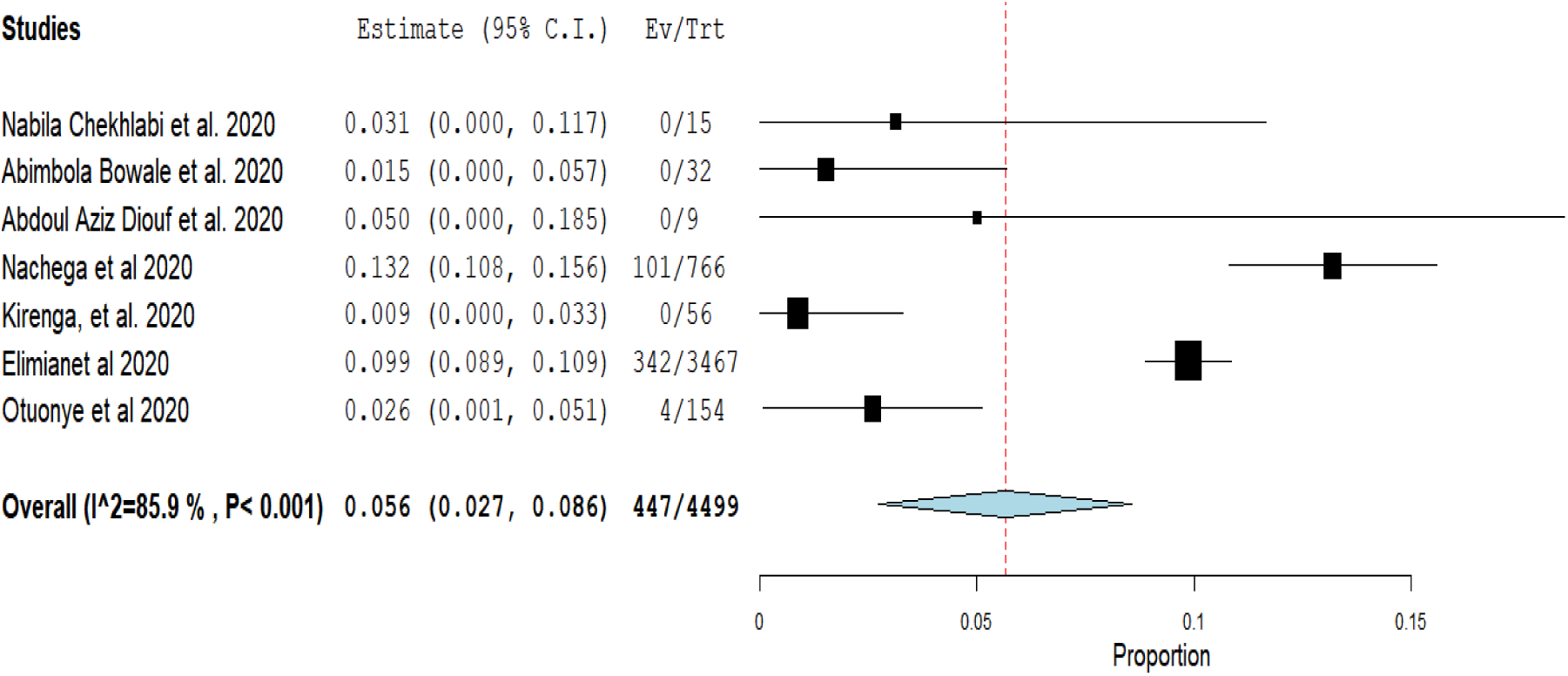
Proportion of death rate in COVID-19 patients examined.

## DISCUSSION

Severe Acute Respiratory Syndrome Coronavirus 2 (SARS-COV-2) is a positive-stranded single-stranded RNA virus within the β-coronavirus cluster^11^. Humans have experienced two prior outbreaks of coronavirus infections - Severe Acute Respiratory Syndrome (SARS) in 2002, and Middle East Respiratory Syndrome (MERS) in 2012, all within the past 2 decades^23^. Since the outbreak of COVID-19 in china, and the first infection record on African soil, identifying the clinical characteristics and its dynamics in COVID-19 patients has been the key effort of most clinical studies. In this study, we evaluated evidence from previous studies in Africa and conducted this meta-analysis to systematically review the clinical characteristics of COVID-19 patients on the continent between June 1 and October 2, 2020. Our analysis included 4490 COVID-19 patients from seven studies conducted in five countries (Morocco, Senegal, Uganda, Congo and Nigeria). To the best of our knowledge,this is the first description and analysis of the clinical characteristics of COVID-19 patients in Africa.

From our analysis, males were the most affected subjects. This finding agrees with the finding of several studies conducted on other continents such as China^1,10,11,13,14^, Kuwait^7^, and the United States^12^. Although, during the SARS-CoV pandemic in 2002, there was a female dominance^24^, and in a study of 869 patients confirmed with COVID-19 in Wuhan, China, there were less men (43.4%)^13^. The most common symptoms of patients with COVID-19 encountered were fever, cough, breathing problem and headache, while diarrhea and rhinorrhea were least encountered. Fever as a clinical symptom of SARS-CoV-2 infection has been found to be associated with other previous coronaviruses infections as documented by SARS-CoV, MERS-CoV, and SARS-CoV-2^24,25^. Our findings of fever, cough and breathing problems as clinical symptoms of COVID-19 were in tandem with the findings of other studies, with up to 99% of patients experiencing fever^24^. However, the proportion of patients that developed fever in our analysis was lower than the reported from meta-analysis done in China and other countries where over 80% of the patients examined had fever^1,10,12,26^. Our study also reported low proportion in cough and breathing problems as compared with studies done outside Africa^11,12^. The case fatality rate from the analyzed data for COVID-19 patients in Africa was lower than reported in other studies,^10,12,14^ but was higher than reports reviewed by Zhu *et al*.^24^ and Rodriguez-Morales *et al*.^27^

This study is not without its strengths and weaknesses. The strengths of this study include its large sample size and the high-quality score of studies included in the analysis. Some limitations were also conducting our meta-analysis. First, most studies included studies were retrospective and conducted at single centers. This may have introduced admission bias and selection bias, so we cannot rule out the influence of other confounding factors. The sample sizes for two studies^9,18^ were relatively small, so the test efficiency may be insufficient. Also, data collected in most of the included studies did not include laboratory findings, hence, it was difficult to analyze the clinicopathological characteristics of the disease in patients across Africa. This meta-analysis showed a significant heterogeneity between the studies, due to too many outcomes, and there was no subgroup analysis, which could affect the accuracy of the results presented.

## CONCLUSION

Currently, documented evidence shows that the most commonly encountered symptoms of COVID-19 patients in Africa were fever, cough, breathing problem and headache. The case-fatality was low, contrary to predictions and modeling results by experts. More high-quality prospective studies are required to verify the above conclusions and to gain more insights into the clinical characteristics of COVID-19 patients in Africa. However, in the absence of any vaccine, relevant measures such as those that address infectious disease outbreak response^28^ should be put in place to slow down the spread of the virus.

## Data Availability

Besides those presented in the study, there are no other supporting data for this manuscript.

## Consent for Publishing

All authors gave their consent for publishing

## Availability of Supporting Data

Besides those presented in the study, there are no other supporting data for this manuscript.

## Competing Interests

All authors declare no competing interests.

## Funding

None

## Authors’ contributions

TJO and LIU were both involved in the conceptualization, design, analysis, and writing of the manuscript. Both authors approved the final draft of the manuscript.

## Notes

### Competing Interest Statement

The authors have declared no competing interest.

### Funding Statement

No external funding was received for this study.

### Author Declarations

No ethical review was necessary for the study (a meta-analysis).

